# From Protocol to Analysis Plan: Development and Validation of a Large Language Model Pipeline for Statistical Analysis Plan Generation using Artificial Intelligence (SAPAI)

**DOI:** 10.64898/2026.03.19.26348626

**Authors:** Hassan Jafari, Petrina Chu, Maximin Lange, Francesca Maher, Callum Glen, Oliver Pearson, Ciel Burges, Louise MacGregor, Meredith Martyn, Samantha Cross, Ben Carter, Richard Emsley, Gordon Forbes

**Author notes:** Corresponding author Maximin Lange, Address IoPPN, 16 De Crispingy Park, SE5, London.

## Abstract

**Background:** Statistical Analysis Plans (SAPs) are essential for trial transparency and credibility but are resource-intensive to produce. While Large Language Models (LLMs) have shown promise in drafting protocols, their ability to generate high-quality, protocol-compliant SAPs remains untested against current content guidance. This study developed and validated an LLM-based pipeline for drafting SAPs from clinical trial protocols.

**Methods:** We developed a structured, section-by-section prompting pipeline aligned with standard SAP guidance. We applied this pipeline to nine clinical trial protocols using three leading LLMs: OpenAI GPT-5, Anthropic Claude Sonnet 4, and Google Gemini 2.5 Pro. The resulting 27 SAPs were evaluated against a 46-item quality checklist derived from the published SAP guidelines. Items were double-scored by independent trial statisticians on a 0–3 scale for accuracy. We compared performance across LLMs and between item types (descriptive vs. statistical reasoning) using mixed-effects logistic regression.

**Results:** Across 9 trials, the models produced SAP drafts with high overall accuracy (77%–78%), with no difference in performance between the three LLMs (p=0.79) but varied by content type (p < 0.001). All models performed well on descriptive items (e.g., administrative details, trial design), with lower accuracy for items requiring statistical reasoning (e.g., modelling strategies, sensitivity analyses). Accuracy for statistical items ranged from 67% to 72%, whereas descriptive items achieved 81% to 83% accuracy. Qualitatively, models were prone to specific failure modes in complex sections, such as omitting necessary details for secondary outcome models or hallucinating sensitivity analyses.

**Discussion:** Current LLMs can effectively draft portions of SAPs, offering the potential for substantial time savings in trial documentation. However, a human-in-the-loop approach remains mandatory; while models demonstrate strong capability in producing descriptive content, their independent application to complex statistical methodology design still requires further methodological development and training. Future work should explore advanced prompt engineering, such as retrieval-augmented generation or agentic workflows, to improve reasoning capabilities.

## Introduction

Artificial Intelligence (AI), and in particular large language models (LLMs), have the potential to accelerate health research by automating the work of researchers, reducing the resources and time needed to conduct research, with potential to accelerate the development and evaluation of new, potentially life changing, interventions. Adoption of AI technology may reduce the time and cost to develop new drugs by up to 50% [1].

Increased levels of research outputs do not necessarily lead to better patient outcomes however, and there is a risk that the adoption of AI in clinical research will reignite ‘the scandal of poor medical research’, with a proliferation of ‘AI slop’: poorly conducted studies executed and reported quickly with help from AI [2]. It is therefore imperative that applications of AI are transparently validated against established methodological standards [3]. In this paper we report on the development of a tool to draft statistical analysis plans for randomised trials, and the evaluation of the tool for statistical accuracy against published guidelines [4].

Statistical analysis plans (SAPs) are central to the credibility of randomised clinical trials. They operationalise the protocol’s scientific objectives into a pre-specified, reproducible analysis strategy. A high-quality SAP helps to: (i) eliminate discretionary analytic decisions made after seeing the data; (ii) provide a technical specification for the coding of trial statistical analysis; and (iii) support transparent reporting and interpretation. Contemporary guidance and regulations therefore position SAPs as a critical companion to the trial protocol and registration [5], providing details on analysis populations, model specifications, multiplicity, missing data methods, and sensitivity analyses [4].

These requirements make SAP authoring both cognitively demanding and time-consuming, particularly given that several sections repeat information from other parts of the SAP and the study protocol. Contemporary regulations require statistical analysis plans to be written under strict operational timelines, prior to the analysis of unblinded data, typically prior to the first Data Monitoring Committee meeting [6]. There is therefore a role for large language models to increase the efficiency in which statistical analysis plans can be written, whilst maintaining or improving the quality of pre-specification.

Large language models (LLMs) have emerged as general-purpose generative AI systems capable of producing coherent, domain-adapted text and structured outputs [7]. Potential roles for LLMs have been suggested in drafting protocol sections, accelerating evidence synthesis workflows, and supporting systematic review tasks such as screening and data extraction, often with human-in-the-loop oversight [2, 8, 9].

A recent review of automation in clinical trial statistical programming provides further context for the potential for the role of LLMs in statistical analysis planning [3]. The review identified nondeterministic output generation, hallucinations, gaps in domain specific knowledge, and a lack of regulatory guidance specific to AI assisted workflows as persistent barriers to AI adoption in clinical trial programming.

In contrast to the growing literature on LLM assistance for protocols, systematic reviews, and statistical programming, there is limited work focused specifically on writing statistical analysis plans. One published case report demonstrated that iterative querying of ChatGPT could yield a plausible SAP for a veterinary randomised trial [10]. More recently, a study has explored the use of LLMs for SAP generation in clinical trials; however, this work did not include formal validation against established methodological guidance or independent expert scoring [11]. The broader question, whether statistical analysis plans can be generated reliably from trial protocols, remains unanswered.

In this study, we developed and validated an AI-based pipeline for drafting SAPs from real-world clinical trial protocols. We evaluated the statistical accuracy and completeness of the generated SAPs, with the aim of quantifying model performance, and providing pragmatic methodological recommendations for the safe and reproducible use of LLMs in SAP authoring.

## Methods

This article is reported following the TRIPOD-LLM reporting guideline [12].

### Prompt development process

Prompt development was conducted iteratively to create a prompt set capable of generating implementable SAP text while minimising irrelevant content. Prompts were developed using four pilot protocols which were not used in the validation. The structure of the prompt library was aligned to the minimum SAP content framework described by Gamble et al. [4], which outlines core SAP sections and item-level expectations.

We used two development stages:

### Stage 1: Vanilla model prompting

The model was provided with the protocol and asked to generate a general SAP (or large SAP portions) with minimal constraints. This served as a baseline to characterise common limitations when using LLMs for SAP drafting. Across protocols, this approach frequently produced outputs that were incomplete with respect to expected SAP elements, variably structured across runs and often mixed content from different parts of the SAP (for example, it would describe study objectives and then immediately move into analysis methods in the same section, making the output difficult to map to standard SAP headings). There were further practical output limits (in particular, response-length/token limits), resulting in insufficiently detailed SAPs. These observations led to a more controlled prompting approach mapped explicitly to SAP guidance.

### Stage 2: Structured, section-by-section prompt library aligned to SAP guidance

To address the issues encountered in stage 1, we developed a structured, modular prompt set that generates the SAP section by section, with one prompt per section. The prompts were designed to correspond directly to the SAP content framework in the Gamble guidance.

Prompts for sections included the following components:

### Common System Message

A common system was used across all sections to standardise the model role (expert clinical trial statistician), specify the protocol as the sole source, and constrain outputs to content appropriate for the requested SAP section. Full prompt instructions can be found on our GitHub repository (https://github.com/rct-sap-ai/sapai-jama-sap-validation).

#### Scope control (“what to include”)

The exact elements required within the section (e.g., randomisation method and stratification factors; sample size assumptions; definition and timing of outcomes).

#### Exclusion rules (“what not to include”)

Prompts explicitly instructed the model to avoid content outside the section remit (e.g., excluding objectives from Study Design; excluding health economics/cost utility analyses from outcome and inferential analysis sections).

#### Protocol-faithfulness instructions

Prompts for areas describing trial design required that the information reported in the SAP was limited to what was stated in the protocol, to reduce unsupported additions or hallucinations.

#### Standardised formatting

Prompts required paragraph-format outputs by default and restricted bullet points to sections where listing was necessary.

#### Handling of non-applicability

Where protocol content could be absent (e.g., interim analyses), prompts instructed the model to make an explicit “not planned” statement rather than extrapolating.

#### Use of examples

To improve clarity and reduce ambiguity, a subset of prompts included brief instructional exemplars demonstrating the expected level of specificity and structure (e.g., how to describe randomisation, how to write outcome definitions as separate paragraphs, or how to express missing data assumptions and permitted methods). This prompting strategy can be described as few shot prompting [13].

The resulting final prompt library covered the main SAP components described in the Gamble et al., guidance, including introduction, study methods (design, randomisation, sample size, interim analyses/internal pilot, timing), statistical principles (significance level, multiplicity, adherence, protocol deviations, analysis populations), trial population, and analysis sections (outcome definitions, primary/secondary analyses, assumptions/sensitivity analyses, subgroup analyses, missing data, additional analyses, and harms).

### Validation and scoring of SAPAI

#### SAP generation for validation

Validation of the SAP generation pipeline was conducted using a convenience sample of nine clinical trial protocols from trials being carried out with the involvement of authors of this study. For each protocol, we generated three SAPs, one per LLM provider/model, using our final prompt pipeline: OpenAI gpt-5 (gpt-5-2025-08-07); Anthropic Claude Sonnet 4 (claude-sonnet-4-20250514); and Google Gemini 2.5 Pro (gemini-2.5-pro). This produced 27 SAPs (9 protocols × 3 models) for evaluation. Prompts were sent using the model’s API, for Google Gemini and Claude Sonnet 4 temperature was set to 0.2 to minimise the variability in output. Default verbosity and reasoning settings were used for OpenAI gpt-5.

#### Validation items and scoring

Validation of the generated SAPs was performed for 33 of the 55 items in the Gamble guidance, excluding items relating to SAP version, SAP revisions, signatures, statistical software or references. Additional validation items were added for the specification of analysis models (items 27a and 27b in Gamble guidance) to align with the PreSPEC framework, covering analysis population, model and covariates. These items were scored twice, once for the primary analysis and once for all secondary analysis.

Each item was rated on an ordinal 4-point scale (0–3):

**0:** item not covered.

**1:** item present but contains substantial omissions, inaccuracies, or hallucinations that could affect scientific integrity if retained in a SAP.

**2:** item present and largely accurate, but with minor omissions/inaccuracies that would not compromise scientific integrity.

**3:** item accurate, protocol-consistent, scientifically appropriate, and (where relevant) sufficiently detailed to allow implementation without ambiguity.

#### Double scoring and independence

Each AI-generated SAP was double-scored by two raters: (1) an experienced trial statistician familiar with the specific protocol (typically the statistician associated with that trial), and (2) an independent trial statistician not involved in that protocol. Raters scored each of the 46 items independently, using a shared scoring guide to promote consistent interpretation across models and trial types. Discrepancies were reviewed item-by-item and resolved via consensus discussions.

### Statistical analysis

#### Statistical analysis of scoring outcomes

The statistical analysis followed a human generated analysis plan, pre-registered prior to generating the validation SAPs [14]. Analysis was conducted using descriptive statistics and inferential results using statistical models. Inferential analyses were carried out using a binary endpoint:

*Scored accurately* = 1 if the item received a score of 3, and 0 if the item received a score of 0, 1, or 2.

A binary endpoint was used to avoid additional model assumptions required when modelling ordinal outcomes

Items were classified a priori into two broad item types:

1. **Descriptive Items**: items that primarily involve reproducing or summarising protocol content (e.g., administrative details, design descriptors, eligibility criteria, timing of assessments), and
2. **Statistical Items:** items that require statistical reasoning and methodological specification beyond simple extraction (e.g., modelling choices, missing data handling, sensitivity analyses).

#### Primary hypothesis: accuracy differs by AI model

The primary hypothesis was that the probability of an item being scored accurately differs across the three LLMs. We tested this using a mixed-effects logistic regression model [15] with outcome: *Scored accurately*, indicator variables for AI model (OpenAI gpt-5, Claude Sonnet 4, Gemini 2.5 Pro) as fixed effects, and random intercepts for trial and item to account for scores for the same item to be correlated, and correlations between scores on the same trial. The pre-specified analysis plan omitted random intercept for item, which was added to ensure clustering in the data was fully taken into account. To test the primary hypothesis, we conducted a single global Wald test assessing whether accuracy differs across models. The primary inference threshold is p < 0.05 (two-sided). A sensitivity analysis was conducted using a mixed effects ordinal model, with the same structure, using the four-point ordinal scale used for scoring.

#### Secondary hypothesis: model performance differs by item type

To assess whether each model performs differently on protocol summarisation items versus statistical reasoning items, we extended the primary mixed-effects logistic model to include item type (statistical item or descriptive item) and a model × item type interaction. An overall test for interaction (evidence that the difference between item types varies by model), and within-model contrasts testing whether accuracy differs by item type for each model were carried out.

As a sensitivity analysis, the primary and secondary analysis were repeated using an ordinal mixed effects model to assess whether outcome dichotomisation affected results.

#### Missing data, exclusions, and software

There were no missing data or exclusions. Statistical analyses were performed using Stata 19.5 [16].

## Results

Table 1 summarises the trials used for the validation.

**Table 1:** Trial Characteristics.

| Trial Acronym | ISRCTN or link to published protocol or results | Randomisation level | Status | Target population | Intervention | Comparator | Primary outcome |
| --- | --- | --- | --- | --- | --- | --- | --- |
| ACTISSIST [17] | ISRCTN - ISRCTN76986679: Active Assistance for Psychological Therapy 2.0 (Actissist 2.0)<br>Results: Effects of Actissist, a digital health intervention for early psychosis: A randomized clinical trial - ScienceDirect | Individuals (1:1), stratified by service (early intervention vs community mental health) | Completed (March 2018 to June 2020) | Adults (16+) with early psychosis (within 5 years of initial episode who are in contact with services) | Actissist app plus usual care | Symptom monitoring app (ClinTouch) plus usual care | Psychotic symptoms measured by Positive and Negative Syndrome Scale total score |
| ATTEND | ISRCTN - ISRCTN17687131: Is an adapted form of mindfulness-based cognitive therapy an effective treatment to support the recovery of 15-18-year-olds who, despite already receiving some treatment, are still experiencing symptoms of depression? | Individuals (1:1), stratified by site | Ongoing, started August 2024 | Young people (15-18) who have completed at least one evidence-based treatment for anxiety or depression still experiencing symptoms of low mood | Mindfulness for Adolescents and Carers (MAC) group therapy plus usual care | Usual care only | Depressive symptoms measured by Short Moods and Feelings total score fortnightly over 12 months follow-up using an Area Under the difference Curve analysis |
| ATTENS [18] | ISRCTN - ISRCTN82129325: Effects of external trigeminal nerve stimulation in ADHD and mechanisms of action<br>Protocol paper: The efficacy of real versus sham external Trigeminal Nerve Stimulation (eTNS) in youth with Attention-Deficit/Hyperactivity Disorder (ADHD) over 4 weeks: a protocol for a multi-centre, double-blind, randomized, parallel-group, phase IIb study (ATTENS) - PubMed | Individuals (1:1), minimisation by sex, medication, site, and age | Ongoing, started September 2021 | Children and adolescents (8-18) with ADHD | Real external trigeminal nerve stimulation (eTNS) | Sham eTNS | Investigator-assessed parent ratings of ADHD-Rating Scale at 6 months |
| DizAi | ISRCTN - ISRCTN41098310: Improving engagement with mental health interventions among low-income university students in Brazil: A feasibility hybrid type III cluster randomised controlled trial | Universities (1:1:1:1) stratified by administrative dependency (public vs private) | Completed (October 2023 to July 2025) | Low-income university students in Brazil enrolled at participating school | Digital group cognitive behaviour therapy plus 1 of three engagement strategy options (A, B, or A+B) | Digital group cognitive behaviour therapy only | Engagement (session attendance at 3 months) |
| EDEN | ISRCTN26462355 - Ketamine for the treatment of depression with anorexia nervosa | Individuals (1:1), stratified by baseline MADRS score ($\leq 30$, $\geq 31$) | Ongoing started October 2025 | Adults with comorbid severe-enduring anorexia nervosa (SE-AN) and major depressive disorder (MDD) | Ketamine capsules (immediate-release formulation; KET-IR), starting dosage of 60mg, flexibility dosed to a maximum of 180mg 2x a week for 4 weeks | Placebo | To assess the recruitment, assessment response rate and drop-out rate as indicators of feasibility |
| HOME | n/a | Individuals (1:1), stratified by site | Start-up | Adults (18+) with major depressive disorder | Transcranial direct current stimulation (tDCS) plus usual care | Usual care only | Depressive symptoms measured by Montgomery-Asberg Depression Rating Scale total score at 10 weeks |
| PROMISE | [ISRCTN - ISRCTN17863382: A clinical trial to test whether treating patients with liver cirrhosis with capsules containing healthy stool bacteria or a dummy capsule (placebo) will reduce the time it takes to develop an infection resulting in hospital admission](https://www.isrctn.com/ISRCTN17863382) | Individuals (1:1), stratified by site | Ongoing, started November 2021 | Adults (18+) with alcohol-related liver disease, metabolic dysfunction-associated steatotic liver disease, or an overlap of the two. | Faecal microbiota transplantation capsules | Matched placebo capsules | Time to first infection resulting in hospital admission |
| RAPID [19] | https://www.isrctn.com/ISRCTN33079589<br>Protocol paper: Study protocol for an adaptive, multi-arm, multi-stage (MAMS) randomised controlled trial of brief remotely delivered psychosocial interventions for people with serious mental health problems who have experienced a recent suicidal crisis: Remote Approaches to Psychosocial Intervention Delivery (RAPID) - PubMed | Individuals (2:2:3), stratified by site and in favour of usual care | Completed (April 2022 to January 2026) | Adults (16+) with serious mental health problems with recent suicidal thoughts/suicide attempt | 1) Structured peer support, 2) Safety planning approach | Usual care only | Any admission at 6 months |
| REACH-ASD [20] | ISRCTN - ISRCTN45412843: REACH-ASD Trial: A randomised controlled trial of a new workshop programme offering information, empowerment and emotional support to parents of children recently diagnosed with Autism Spectrum Disorder<br>Protocol paper: REACH-ASD: a UK randomised controlled trial of a new post-diagnostic psycho-education and acceptance and commitment therapy programme against treatment-as-usual for improving the mental health and adjustment of caregivers of children recently diagnosed with autism spectrum disorder - PMC | Individuals (2:1), stratified by site and in favour of group therapy | Completed (September 2019 to April 2022) | Parents/primary caregivers of children aged 2-15 who have received a diagnosis of ASD in the last 12 months in the Greater Manchester area | Empower-Autism group programme plus usual care | Local post-diagnostic offer plus usual care (except those similar to Empower-Autism) | Parental mental health as measured by the General Health Questionnaire 30-item total score at 52 weeks |
| STAR [21] | ISRCTN - ISRCTN93382525: STAR (Study of Trauma And Recovery): a trial of trauma-focused psychological therapy for psychosis<br>Protocol paper: Multisite randomised controlled trial of trauma-focused cognitive behaviour therapy for psychosis to reduce post-traumatic stress symptoms in people with co-morbid post-traumatic stress disorder and psychosis, compared to treatment as usual: study protocol for the STAR (Study of Trauma And Recovery) trial - PubMed | Individuals (1:1), stratified by site | Completed (February 2020 to February 2025) | Adults (18+) with PTSD and schizophrenia spectrum diagnoses from five NHS mental health trusts | Trauma-focused cognitive behaviour therapy for psychosis plus usual care | Usual care only | PTSD symptoms as measured on Clinician-Administered PTSD Scale for DSM-5 at 9 months |

### Inferential results

Table 2 shows the results of statistical tests for comparisons between models, and between items relating to statistical modelling and descriptive items. There was no evidence of differences between models in the proportions of items scored as being correct, either overall, for items relating to statistical modelling, or descriptive items. All models performed substantially worse for items relating to statistical modelling compared to descriptive items, 72% vs 81% for GPT5 respectively, 67% vs 83% for Gemini Pro 2.5, and 70% vs 83% for Claude Sonnet. The sensitivity analysis using an ordinal mixed effects model found similar results.

Figure 1 shows performance by model by section of the SAP. This figure shows that the models do extremely well on sections of the SAP which involve administrative information or describing the trial and less well in areas relating to the analysis or outcome definitions. For example, Claude Sonnet picked up on a sample size recalculation for a randomised trial with clustering in intervention arm only and summarised the changes in assumptions neatly:

**Figure 1.**
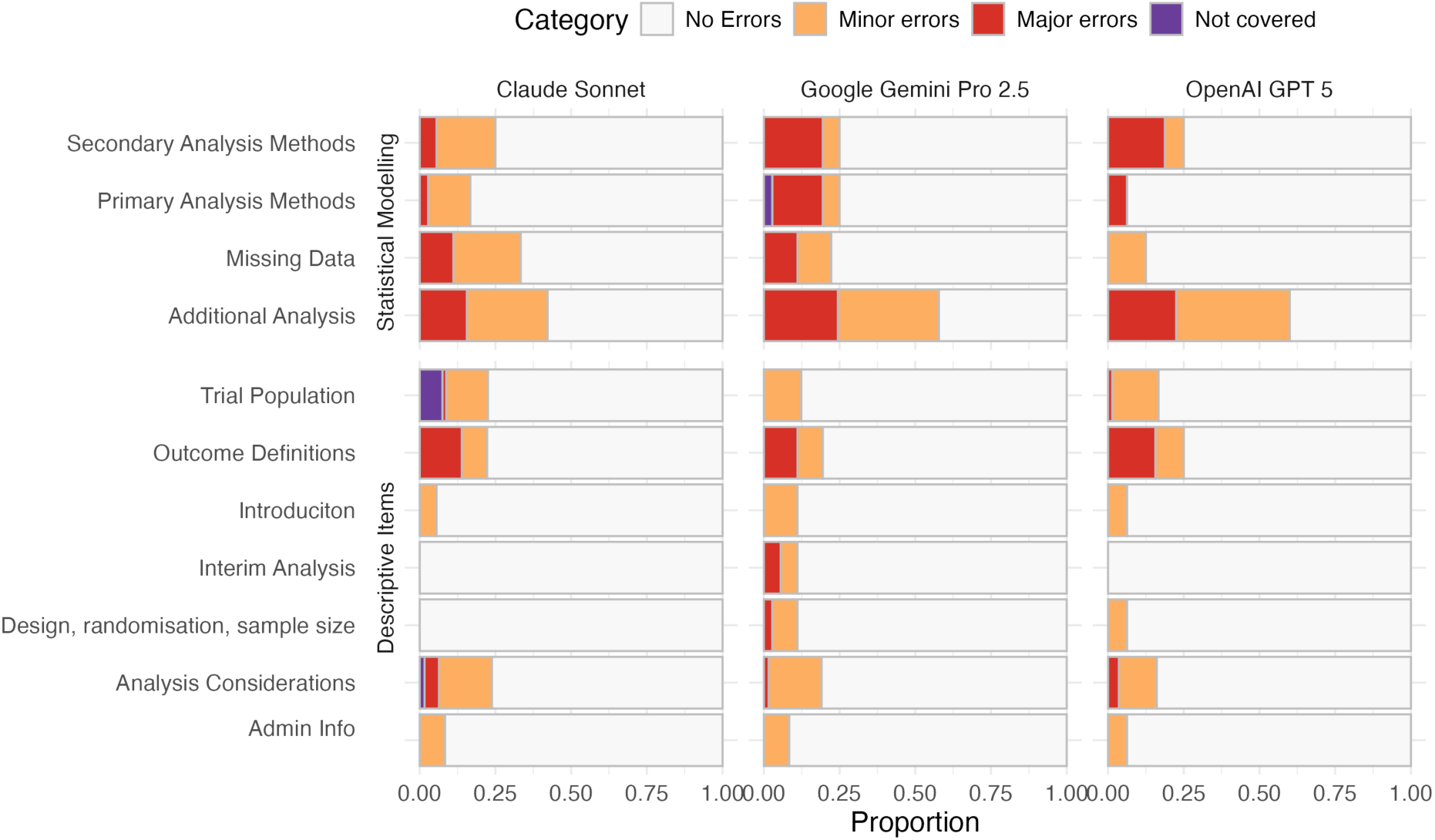
Results by model and section of the statistical analysis plan.

**Table 2:**
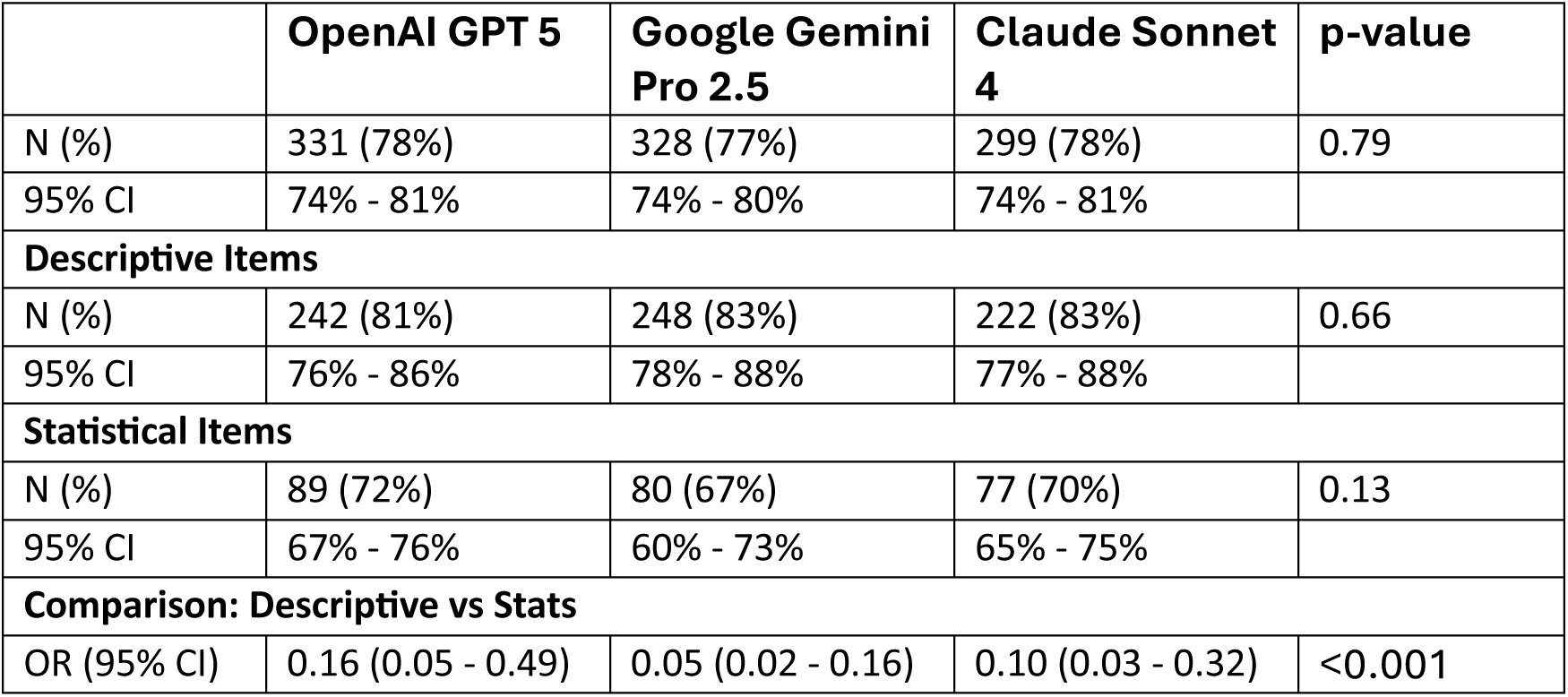
Comparison of accuracy between models, and between items directly relating to statistical modelling and descriptive items. The table shows the number and proportion of items scored as correct for each model over all trials. Proportions and 95% CIs are taken from marginal model predictions. The final column, global p-value shows the result of a test of all models having equal performance. For the descriptive vs stats comparison the final column shows the results comparing descriptive and statistical items, assuming all models have the same performance.

> The revised calculation assumed intervention groups of average size 8 with variation in group size of 8, maintaining an intracluster correlation coefficient of 0.02 in the treatment arm. The attrition rate was increased to a more conservative 25% across both arms. (REACH-ASD)

Whilst there were more errors amongst items requiring statistical reasoning there were some good examples. In this example Claude Sonnet correctly described a shared frailty cox regression model for a time to event primary outcome:

> The primary outcome of time to first infection resulting in hospital admission will be analysed using a Cox proportional hazards model with frailty effect to account for site heterogeneity. The model will include treatment group as the main effect and adjust for baseline disease severity, participant age, sex, and baseline cirrhosis severity as covariates. The treatment effect will be presented as a hazard ratio with S5% confidence interval. Proportionality will be assessed using Kaplan-Meier survival and log-log plots. (PROMISE, Claude Sonnet).

Selected examples of errors and scores are given in Table 3. The errors described for the outcomes are simple omissions that may be corrected by careful review. Errors in the statistical analysis methods or sensitivity analysis are more subtle, the proposed could look reasonable, however are not the optimal methods and could lead to lower precision of estimates, wider confidence intervals, and potentially materially change the interpretation of the trial.

**Table 3.** Examples of errors, the score given, and a justification.

| **SAP section** | **Error details** | **Score** |
| --- | --- | --- |
| Sample size | In a multi-arm multi-stage trial protocol, the overall family-wise error rate (FWER) was incorrectly stated as being controlled at 0.0403 when the protocol states that the overall FWER for the trial is controlled at 0.0397. Later in the analysis section the model stated an error rate of 0.05, as is conventionally used. | 2 = Minor Error as a small mistake when describing the error rate would not impact the conclusions of the trial analysis. |
| Outcome definitions | Models sometimes missed outcomes, or incorrectly classified time-to-event outcomes as counts or rates rather than time to event. | 1 = Major Error as omission of outcomes from an analysis pre-specification would lead to findings not being treated as confirmatory. Treating outcomes as counts leads to an incorrect analysis approach. |
| Primary analysis | Models frequently missed covariates when specifying analysis that were included in the protocol. | 1 = Major Error<br>Omitting important covariates could reduce the precision of estimates, potentially changing trial conclusions. |
| Secondary analysis | The models proposed linear regression models for the analysis of continuous outcomes assessed at multiple post follow up timepoints. Repeated measures mixed models would have been more efficient. | 1 = Major Error<br>Not making use of the most efficient analysis model could reduce the precision of estimates, potentially changing trial conclusions. |
| Sensitivity analyses | Proposed sensitivity analysis frequently included plausible analysis that would not necessarily be appropriate.<br>This included proposed per-protocol analyses, checks for non-linearity of associations between baseline covariates and outcome, with modifications to the reported primary analysis if non-linearities identified. | 1 = Major Error<br>Conducting unnecessary, and potentially biased sensitivity analysis could change the interpretation of the primary analysis result. |

## Discussion

Our findings demonstrate that current LLMs can generate high-quality first drafts of SAPs with reasonable accuracy. Importantly, we found no difference in performance between the three model providers, suggesting that the utility of LLMs for this task is becoming a generalisable capability of state-of-the-art models rather than a feature specific to a single proprietary architecture. To our knowledge, this study represents the first validation of a LLM pipeline designed specifically for drafting Statistical Analysis Plans for randomised trials.

However, the high overall accuracy masks a critical dichotomy in performance. While models excelled at extraction-based tasks on routine sections such as administrative details and trial design, their performance dropped on items requiring statistical reasoning, such as model specification and sensitivity analyses. This points to a caveat for the safe adoption of AI in clinical trial units: LLMs function effectively as efficient technical writers for descriptive content but currently lack the reliability to act as autonomous statistical architects.

The observed performance gap between routine and statistically in-depth items aligns with the broader literature on LLM capabilities in specialised domains [3]. The models proved highly effective at translating protocol text into the structured format required by the Gamble et al. guidelines [4].

This capability offers immediate value by automating the substantial administrative and descriptive boilerplate that constitutes a large portion of a standard SAP. By reducing the time required to draft these sections, trial statisticians can redirect cognitive effort toward the complex methodological challenges where human statistical expertise remains essential.

Our validation study highlights another risk with relying solely on LLMs: the models frequently struggled with correctly specifying all secondary outcome analysis and struggled with the nuance of sensitivity analyses which whilst on the surface appeared valid, would not be appropriate analyses for randomised trials. These plausible hallucinations, in which a text reads professionally but is methodologically incorrect, are more dangerous than easy to spot mistakes. A model might convincingly describe a mixed-effects model for a secondary outcome that differs subtly but materially from the primary analysis strategy or fabricate a tipping point analysis that the investigators never intended to perform. Consequently, these tools must be viewed strictly as human-in-the-loop drafting assistants. The generated SAP should be treated as a template requiring comprehensive validation, particularly for sections involving inferential logic and estimand definition.

A strength of this study is the robust validation framework. Unlike previous case reports that relied on subjective assessment of single outputs [10] or utilised a brief partly-automated evaluation framework [11], we employed a double-blind scoring system using independent trial statisticians and a standardised checklist derived from JAMA guidance [4]. Furthermore, by testing across three distinct model families, we provide evidence that is robust to the rapidly changing landscape of AI vendors.

However, several limitations must be acknowledged. First, our sample size was limited to nine protocols. Second, our prompting strategy, while structured, relied on a one-shot or few-shot approach per section. We did not employ iterative feedback loops where the model critiques its own output, which might have improved reasoning performance. Thirdly, the validation was conducted on the generated text as is in practice, a statistician would likely interact with the model to refine the output, potentially achieving higher final quality than measured here. Current analysis plan writing processes typically involve review by multiple statisticians, and issues around ambiguous specification of statistical methods still remain. Finally, we relied on published guidance for the validation which does not reflect the latest methodological advances, in particular it does not include any reference to the estimands framework [22–24].

The current pipeline establishes a baseline for automated SAP generation, but future work should focus on improving statistical accuracy. Potential technical avenues could include a retrieval-augmented generation architecture, where models have access to external statistical resources or guidelines, as well as agentic systems in which one AI agent drafts the SAP and another acts as a reviewer to check against the protocol. This could mimic the human quality assurance process and catch errors before the human user sees the draft, and integrate SAP generation and protocol development. If LLMs are used to help draft the protocol, they could simultaneously generate the corresponding SAP sections, ensuring potentially perfect alignment between the two documents from the outset.

Whilst this study has focused on writing statistical analysis plans for randomised trials, it has implications beyond this setting. Randomised trials offer a relatively benign challenge due to their highly structured and regulated conduct. We would expect the challenges in accuracy reported in this study to be as great or greater if LLMs were used to plan the analysis for observational data, where the range of research questions, and methods used to address them is far broader. Secondly, the prompting strategy we used is similar to the approach an individual would take when informally querying the models about a statistical analysis. Our findings are therefore also potentially relevant to the general use of LLMs to aid statistical analysis.

This study confirms that AI-assisted SAP authoring is not only feasible but highly effective for large portions of the document. A simple, structured prompting approach can produce drafts that are mostly accurate, offering the potential for substantial efficiency gains in clinical trial operations. However, the specific deficits in statistical reasoning underscore the enduring need for expert statistical oversight and the widespread use of AI to plan statistical analysis by researchers without sufficient expertise will jeopardise research integrity. LLMs are powerful accelerators for the trial statistician, but they remain, for now, the draughtsman rather than the architect.

## Data Availability

SAP-AI is live and free to use for anyone at https://sapai-streamlit-production.up.railway.app/

Source code for SAP-AI can be found at https://github.com/rct-sap-ai/sap-kcl

Source Code for all analysis reported can be found at https://github.com/rct-sap-ai/validation-study-analysis/

